# Analytical and clinical performances of five immunoassays for the detection of SARS-CoV-2 antibodies in comparison with neutralization activity

**DOI:** 10.1101/2020.08.01.20166546

**Authors:** Andrea Padoan, Francesco Bonfante, Matteo Pagliari, Alessio Bortolami, Davide Negrini, Silvia Zuin, Dania Bozzato, Chiara Cosma, Laura Sciacovelli, Mario Plebani

**Affiliations:** Department of Medicine-DIMED, Medical School, University of Padova, Italy; Department of Laboratory Medicine, University-Hospital of Padova, Italy; Laboratory of Experimental Animal Models, Division of Comparative Biomedical Sciences, Istituto Zooprofilattico Sperimentale delle Venezie, Legnaro, Italy

**Keywords:** SARS-CoV-2, immunoassays, serology, antibodies, clinical performances

## Abstract

**Background:** Reliable high-throughput serological assays for SARS-CoV-2 antibodies (Abs) are urgently needed for the effective containment of the COVID-19 pandemic, as it is of crucial importance to understand the strength and duration of immunity after infection, and to make informed decisions concerning the activation or discontinuation of physical distancing restrictions.

**Methods:** In 184 serum samples from 130 COVID-19 patients and 54 SARS-CoV-2 negative subjects, the analytical and clinical performances of four commercially available chemiluminescent assays (Abbott SARS-Cov-2 IgG, Roche Elecsys anti-SARS-CoV-2, Ortho SARS-CoV-2 total and IgG) and one enzyme-linked immunosorbent assay (Diesse ENZY-WELL SARS-CoV-2 IgG) were evaluated and compared with the neutralization activity achieved using the plaque reduction neutralization test (PRNT).

**Findings:** Precision results ranged from 0.9% to 11.8% for all assays. Elecsys anti-SARS-CoV-2 demonstrated linearity of results at concentrations within the cut-off value. Overall, sensitivity ranged from 78.5 to 87.8%, and specificity, from 97.6 to 100%. On limiting the analysis to samples collected 12 days after onset of symptoms, the sensitivity of all assays increased, the highest value (95.2%) being obtained with VITRO Anti-SARS-CoV-2 Total and Architect SARS-CoV-2 IgG. The strongest PRNT_50_ correlation with antibody levels was obtained with ENZY-Well SARS-CoV-2 IgG (rho = 0.541, p < 0.001).

**Interpretation:** The results confirmed that all immunoassays had an excellent specificity, whereas sensitivity varied across immunoassays, depending strongly on the time interval between symptoms onset and sample collection. Further studies should be conducted to achieve a stronger correlation between antibody measurement and PRNT_50_ in order to obtain useful information for providing effective passive antibody therapy, and developing a vaccine against the SARS-CoV-2 virus.

## Introduction

The continuing spread of coronavirus disease 2019 (COVID-19) caused by severe acute respiratory syndrome coronavirus 2 (SARS-CoV-2) has prompted concern worldwide, leading the World Health Organization (WHO) to declare COVID-19 a pandemic on 11 March 2020 ^1^. The accurate and timely diagnosis of the disease is crucial to the effective management of patients, control of the pandemic and the establishment of appropriate infection control measures. Although real-time reverse transcription polymerase chain reaction (rRT-PCR) allows the diagnosis of the disease in most patients, including asymptomatic carriers, it has some analytical and clinical limitations. Analytical pitfalls in both the pre- and analytical steps have been described ^2^ and negative molecular test results have been reported in the later stages of infection, thus being misleading from a clinical viewpoint. Therefore, rRT-PCR precludes the identification of individuals who have been infected, but have had only minor, or no, symptoms and therefore have not sought medical attention. A wide range of immunoassays to detect SARS-CoV-2 antibodies (Ab) have been developed to complement rRT-PCR, with different antigen targets and formats ^3–5^. Although not effective for making an early diagnosis, serological assays for SARS-CoV-2 play an important role in diagnosing COVID-19 disease in individuals who present late, in understanding the virus epidemiology in the general population, and in identifying the disease prevalence in categories at higher risk of infection (e.g. healthcare workers). In addition, they should be used to ascertain the efficacy of containment measures both locally and globally, to screen convalescent sera for therapeutic and prophylactic purposes, and to improve knowledge of the immune response to the novel virus as the degree and duration of the response of specific antibodies is as yet poorly understood ^6,7^. Like infections from other pathogens, SARS-CoV-2 infection elicits development of IgM and IgG specific Ab which are the most available antibodies for assessing response, while little is known about IgA response in the blood.

The aim of this paper is to evaluate the performance characteristics and diagnostic specificity, sensitivity of four chemiluminescent assays (CLIA) and one enzyme-linked immunosorbent assay (ELISA) for SARS-CoV-2 antibodies and in comparison with neutralizing activity.

## Material and Methods

### Patients

A total of 184 leftover serum samples from 130 COVID-19 patients (8 asymptomatic/paucisymptomatic who recovered at home with supportive care and isolation, and 122 hospitalized classified with moderate or severe disease, following WHO interim guidance^8^) and 54 SARS-CoV-2 negative subjects (33 healthcare workers, 21 autoimmune patients, 8 pregnant women) were included in the study (Table 1). All subjects underwent at least one nasopharyngeal swab test, analyzed by RT-PCR. Healthcare workers were considered negative (Neg-HW) on the basis of at least three negative sequential molecular test results obtained between February 26th and May 29th, 2020. The study protocol (number 23307) was approved by the Ethics Committee of the University-Hospital, Padova.

**Table 1:** Demographic characteristics of the 184 subjects included in the study.

| Groups | N (%) | Gender |  | Age (mean±SD) |
| --- | --- | --- | --- | --- |
|  |  | Females N (%) | Males N (%) |  |
| **Negative Healthy Workers (Negative)** | 33 (17.9%) | 25 (75.7%) | 8 (24.3%) | 40.1±11.8 |
| **Autoimmune Patients and Pregnant women (AI/Pr)** | 21 (11.4%) | 19 (90.5%) | 2 (9.5%) | 43.8±16.2 |
| **Asymptomatic / Paucisymptomatic SARS-CoV-2 positive Patients (Asympt)** | 8 (4.4%) | 7 (87.5 %) | 1 (12.5%) | 45.4±17.9 |
| **Moderate/Severe SARS-CoV-2 positive patients (SARS-CoV-2)** | 122 (66.3%) | 36 (29.5%) | 86 (70.5%) | 64.5±16.6 |
| **Overall** | 184 (100%) | 87 (47.3%) | 97 (52.7%) | 56.9±19.1 |

### Analytical systems under evaluation

In this study, an evaluation was made of four commercially available chemiluminescent immunoassays (CLIA) (Anti-SARS-CoV-2 IgG and Total, Ortho Clinical Diagnostics, Raritan, NJ, USA; Elecsys Anti-SARS-CoV-2, Roche Diagnostic GmbH, Mannheim, Germany; SARS-CoV-2 IgG, Abbott Laboratories, IL, USA) and one enzyme-linked immunosorbent assay (ELISA) (ENZY-WELL SARS-CoV-2 IgG, Diesse Diagnostica Senese, Siena, Italy). Table 2 provides a summary of the specific features of each immunoassay.

**Table 2:** The five SARS-CoV-2 antibody assays investigated: characteristics specified by the manufacturers.

| Manufacturer | Ortho Clinical Diagnostics |  | Roche Diagnostics | Abbott | DIESSE Diagnostics |
| --- | --- | --- | --- | --- | --- |
| **Commercial name** | Anti-SARS-CoV-2 IgG | SARS-CoV-2 Total | Elecsys Anti-SARS-CoV-2 | SARS-CoV-2 IgG (also referred to as CoV-2 IgG) | ENZY-WELL SARS-CoV-2 IgG |
| **Platform** | VITROS ECi/ECiQ/3600 and VITROS 5600/XT 7600 | VITROS ECi/ECiQ/3600 and VITROS 5600/XT 7600 | Cobas e 411, Cobas e 601 and Cobas e 602 | All Architect systems | 96 wells microplate, automatable |
| **Method** | Chemiluminescent immunoassay (CLIA) | Chemiluminescent immunoassay (CLIA) | Electro-ChemiLuminescent (ECLIA) | Chemiluminescent Microparticle Immunoassay (CMIA) | Enzyme-linked immunosorbent assay (ELISA) |
| **Detection** | IgG Antibodies against SARS-CoV-2 | Total Antibodies (Included IgG, IgA e IgM) | Antibodies (included IgG) against SARS-CoV-2 | IgG antibodies against SARS-CoV-2 | IgG antibodies against SARS-CoV-2 |
| **Antigen target** | Spike Protein | Spike protein S1 | Nucleocapsid protein | Nucleocapsid protein | Native antigen (Vero E6 cells infected with SARS-CoV-2) |
| **Results** | Signal/Cut-off (S/C) | Signal/Cut-off (S/C) | Signal Sample/Cut-off (COI) | Index (S/C) | OD/Cut-off (Index) |
| **Interpretation** | < 1.0 Negative<br>≥ 1.0 Positive | < 1.0 Negative<br>≥ 1.0 Positive | < 1.0 Negative<br>≥ 1.0 Positive | < 1.4 Negative<br>≥ 1.4 Positive | < 0.9 Negative<br>0.9 - 1.1 Equivocal<br>≥ 1.1 Positive |
| **Sensitivity** | 12-15 d: 83.3%<br>≥ 8 d: 90.0% | 0-8 d: 100.0%<br>≥ 8 d: 100.0% | 0-6 d: 65.5%<br>7-13 d: 88.1%<br>≥ 14 d: 100% | < 3 d: 0.0%<br>3-7 d: 25.0%<br>8-13 d: 86.4%<br>≥ 14 d: 100% | 92.5% |
| **Specificity** | 100.0% | 100.0% | 99.80% | Pre-COVID-19: 99.6%<br>Other Respiratory Illness: 100.0% | 95.8% |

d = Days.

Moreover, Liaison SARS-CoV-2 S1/S2 IgG (Diasorin, Sallugia-VC, Italy), ENZY-Well SARS-CoV-2 IgA and IgM were evaluated for the correlation with the neutralization results.

### Repeatability and intermediate precision evaluation

Precision estimation was performed on CLIA assays using two human serum sample pools with different values, by means of quintuplicate measurements of same pool aliquots, performed for a total of four consecutive days. Nested analysis of variance was used to estimate precision, following the CLSI EP15-A3 protocol ^9^ The results for precision were compared to those claimed by the manufacturer when available, using the procedure recommended by EP15-A3. Repeatability and within-laboratory precision were in accordance with the repeatability and intermediate precision conditions specified in the international vocabulary of metrology (VIM, JCGM 100:2012) for precision estimation within a four-day period.

### Linearity assessment

Linearity was assessed using serial dilution of two samples pools, prepared with two different levels of SARS-CoV-2 antibodies (high and low level pools), as specified in the CLSI EP06-A, guideline (paragraph 4.3.1) ^10^. In brief, the following high-level serum pools were prepared: 5.2 signal to cut-off (S/CO) ratio for VITROS Anti-SARS-CoV-2 IgG, 53 S/CO ratio for VITROS Anti-SARS-CoV-2 total, 2.71 S/CO ratio for Elecsys Anti-SARS-CoV-2, and 3.76 S/CO ratio for Architect SARS-CoV-2 IgG. The pools were serially diluted with the corresponding low-level serum pools (0.174 S/CO ratio for Elecsys Anti-SARS-CoV-2, 0.01 S/CO for VITROS Anti-SARS-CoV-2 IgG, 0.2 S/CO for VITROS Anti-SARS-CoV-2 total, 0.04 S/CO ratio for Architect SARS-CoV-2 IgG). All measurements were performed in triplicate.

### Plaque reduction neutralization test (PRNT)

For a subgroup of 52 samples from SARS-CoV-2 positive subjects, the Liaison SARS-CoV-2 S1/S2 IgG (Diasorin, Sallugia, VC, Italy) ^5^, ENZY-WELL SARS-CoV-2 IgA and IgM (Diesse Diagnostica Senese, Siena, Italy) assays were further performed. In the same samples, a high-throughput PRNT method was developed for the fast and accurate quantification of neutralizing antibodies in plasma samples collected from patients exposed to SARS-CoV-2. Samples were heat-inactivated by incubation at 56 °C for 30 minutes and 2-fold dilutions were prepared in Dulbecco modified Eagle medium (DMEM). The dilutions, mixed to a 1:1 ratio with a virus solution containing 20-25 focus-forming units (FFUs) of SARS-CoV-2, were incubated for 1 hour at 37□°C. Fifty microliters of the virus–serum mixtures were added to confluent monolayers of Vero E6 cells, in 96-wells plates and incubated for 1 hour at 37□°C, in a 5% CO_2_ incubator. The inoculum was removed and 100 µl of overlay solution of Minimum essential medium (MEM), 2% fetal bovine serum (FBS), penicillin (100 U/ml), streptomycin (100 U/ml) and 0.8% carboxy methyl cellulose was added to each well. After 26 hours’ incubation, cells were fixed with a 4% paraformaldehyde (PFA) solution. Visualization of plaques was obtained with an immunocytochemical staining method using an anti-dsRNA monoclonal antibody (J2, 1:10,000; Scicons) for 1 hour, followed by 1-hour incubation with peroxidase-labeled goat anti-mouse antibodies (1:1000; DAKO) and a 7-minute incubation with the True Blue™ (KPL) peroxidase substrate. FFUs were counted after acquisition of pictures at a high resolution of 4800 x 9400 dpi, on a flatbed scanner. The serum neutralization titer was defined as the reciprocal of the highest dilution resulting in a reduction of the control plaque count >50% (PRNT_50_). We considered a titer of 1:10 as the seropositive threshold.

### Statistical analyses

For evaluation of precision, an in-house developed R (R Foundation for Statistical Computing, Vienna, Austria) script for implementing the CLSI EP15-A3 protocol was used for ANOVA and for calculating the upper verification limit^9^. The GraphPad Prism version 8.4.1 for Windows (GraphPad Software, LLC) was employed to evaluate plaque reduction neutralization test results, using non-parametric tests (Kruskall-Wallis test and Spearman’s correlation). Stata v13.1 (Statacorp, Lakeway Drive, TX, USA) was used to evaluate the assays’ clinical performances. Bonferroni’s adjusted p-value (B-adj) was calculated for multiple comparisons. For ROC analyses, a/the non-parametric empirical method was used to estimate the area under the ROC curve (AUC), while the ‘diagt’ module was used to estimate sensitivity, specificity, and positive and negative predictive values.

## Results

### Patients’ characteristics

Table 1 reports the demographic characteristics of the subjects included in the study. The overall mean age of subjects was 56.4 years, with a standard deviation (±SD) of 18.7 (range 22.7 - 92.2 years). The ages of negative healthy workers (Neg-HW) [Bonferroni’s adjusted p-value (B-adj) p < 0.0001], autoimmune/pregnant subjects (AI/Pr) (B-adj p < 0.0001) and asymptomatic/paucisymptomatic (Asympt) subjects (B-adj p < 0.0001) differed from that of SARS-CoV-2 patients. The percentage of females differed significantly from that of males (p< 0.001), particularly in the AI/Pr group. For SARS-CoV-2 patients, the mean time interval from the onset of symptoms and serological determinations was 24.4 days (SD ±17.9; range 4 - 89 days).

### Repeatability and intermediate precision

Results for precision of all the CLIA assays are reported in Table 3. The ANOVA approach allowed us to estimate repeatability and intermediate precision separately. Only the Architect SARS-CoV-2 IgG insert reported data on precision, claimed at levels of 0.04 and 3.53 S/CO ratio. For this immunoassay, intermediate precision performances statistically deviated from the manufacturer’s claims at both levels. All the immunoassays had acceptable analytical imprecision (CV%).

**Table 3:** Precision results for the studied immunoassays. Coefficient of variation (CV) are expressed in percentage (%) and were obtained by using pools of samples.

| Measurand | Level | Design | Laboratory evaluation of repeatability, CV % | Laboratory Evaluation of Intermediate precision - CV% |
| --- | --- | --- | --- | --- |
| **Architect SARS-CoV-2 IgG<sup>#</sup>** | 0.05 (ratio S/CO)<sup>#</sup> | 5x4 | 10.3* | 11.8* |
|  | 3.81 (ratio S/CO)<sup>#</sup> |  | 0.9 | 1.75* |
| **VITROS Anti-SARS-CoV-2 IgG<sup>\$</sup>** | 0.01 (ratio S/CO) | 5x4 | < 0.1 | < 0.1 |
|  | 5.35 (ratio S/CO) |  | 4.35 | 4.35 |
| **VITROS Anti-SARS-CoV-2 Total<sup>^</sup>** | 0.03 (ratio S/CO) | 5x4 | 18.30 | 30.51 |
|  | 51.2 (ratio S/CO) |  | 1.97 | 2.82 |
| **Elecsys Anti-SARS-CoV-2<sup>°</sup>** | 0.73 (ratio S/CO) | 5x4 | 1.21 | 2.84 |
|  | 2.57 (ratio S/CO) |  | 0.87 | 2.63 |

<sup>#</sup> obtained from the Abbott Architect insert CoV-2 IgG 6r86, H07891R02, B6R860 revised April 2020.
<sup>\$</sup> obtained from the VITROS Anti-SARS-CoV-2 IgG Ortho Clinical Diagnostic insert v4.0 pub. No. GEM1292\_US\_EN.
<sup>^</sup> obtained from the VITROS Anti-SARS-CoV-2 Total Ortho Clinical Diagnostic insert v4.0 pub. No. GEM1293\_XUS\_IT.
<sup>°</sup> obtained from the Cobas Anti-SARS-CoV-2 Roche Diagnostic insert 09203095500 v1.0, 2020-05 in Italian.
\* indicates that imprecision value was higher than that declared by manufacturers, also after the calculation of UVL as suggested by EP15-A3 (5.4% for Repeatability and 5.9% for Intermediate precision at level 0.04 Index S/C and 1.1 for Repeatability and 1.2% for Intermediate precision at level 3.53 S/CO).

### Linearity assessment

Linearity results for all the CLIA studies are summarized in Figure 1. All tested mixes of sample pools covered a wide range of values and included the manufacturers’ cut-offs. All immunoassays, except for Elecsys Anti-SARS-CoV-2, deviated from linearity, the coefficients of the second-order polynomial fit attaining high statistical significance.

**Figure 1:**
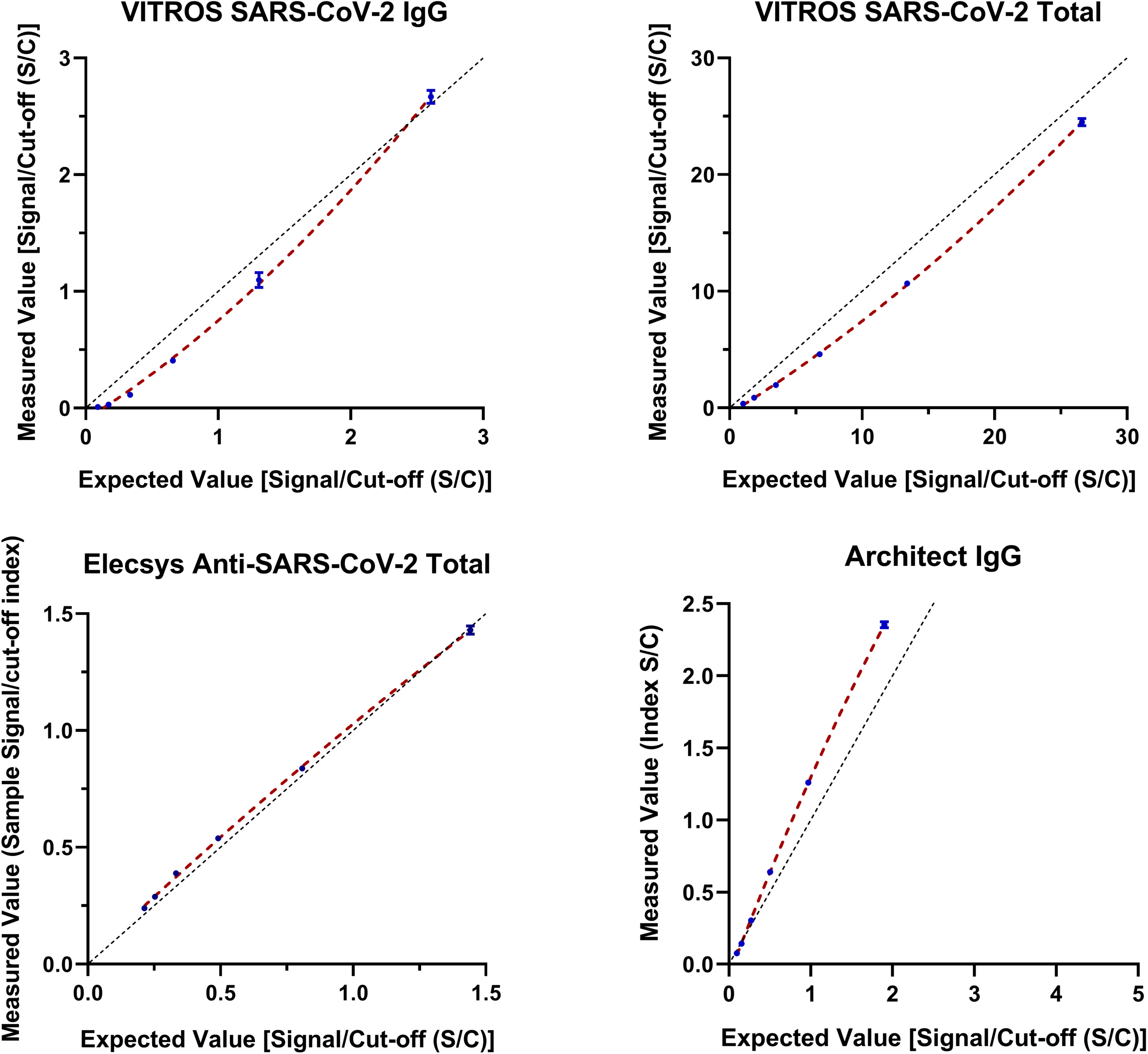
Linearity assessment of the studies immunoassays.

### Evaluation of clinical performances

Sensitivities, specificities, and positive and negative likelihood ratios were estimated using the manufacturers’ cut-offs, while receiver operating characteristic (ROC) curves were used to evaluate overall performance(s). Elecsys Anti-SARS-CoV-2 immunoassay results were available for 172/184 (93.4%) serum samples. Table 4 summarizes estimated clinical performances for all CLIA and the ELISA immunoassays considering the total time frame of 93 days and limiting the analyses to sera collected 12 days after the onset of symptoms. One hundred fifty-eight samples were included and evaluated in this restricted subgroup, while only 146 results were available for Elecsys Anti-SARS-CoV-2. Table 4 shows data on positive and negative likelihood ratios, allowing an easy estimation of positive (PPV) and negative (NPV) predictive values given disease prevalence. Considering two different scenarios of disease prevalence settings: (a) 4%, as found in a Veneto Region (Italy) survey (data not shown) ^11^; (b) 10%, as described in a survey conducted in Geneva ^12^, PPV and NPV were then estimated, using VITROS Anti-SARS-CoV-2 Total and Architect SARS-CoV-2 IgG immunoassays for comparative purposes. Regarding performances calculated 12 days after the onset of symptoms, VITROS Anti-SARS-CoV-2 Total PPV (95%CI) and NPV (95%CI) were 66.3% (22.0-93.2%) and 99.5% (99.2-99.7%) with a prevalence of 4%, 84% (43.0%-97.3%) and 98.6% (97.8%-99.1%) with a prevalence of 10%. Within the total time frame, results for PPV (95%CI) and NPV (95%CI) changed to 68.2% (23.5-93.7%) and 99.8% (99.5-99.9%) with a prevalence of 4%, 85.1% (45.0-97.6%) and 99.5% (98.7-99.8%) with a prevalence of 10%.

Architect SARS-CoV-2 IgG PPV (95%CI) and NPV (95%CI) were 100% (21.6-98.6%) and 99.8% (99.5-99.9%) with a prevalence of 4%, 100% (42.3-99.5%) 99.5% (98.7-99.7%) with a prevalence of 10%. On considering the total time frame, these results changed to 100% (19.7-98.4%) and 99.4% (99-99.6%), and 100.0% (39.5-99.4%) and 98.3% (97.5-98.8%) with prevalence settings of 4% and 10%.

**Table 4:**
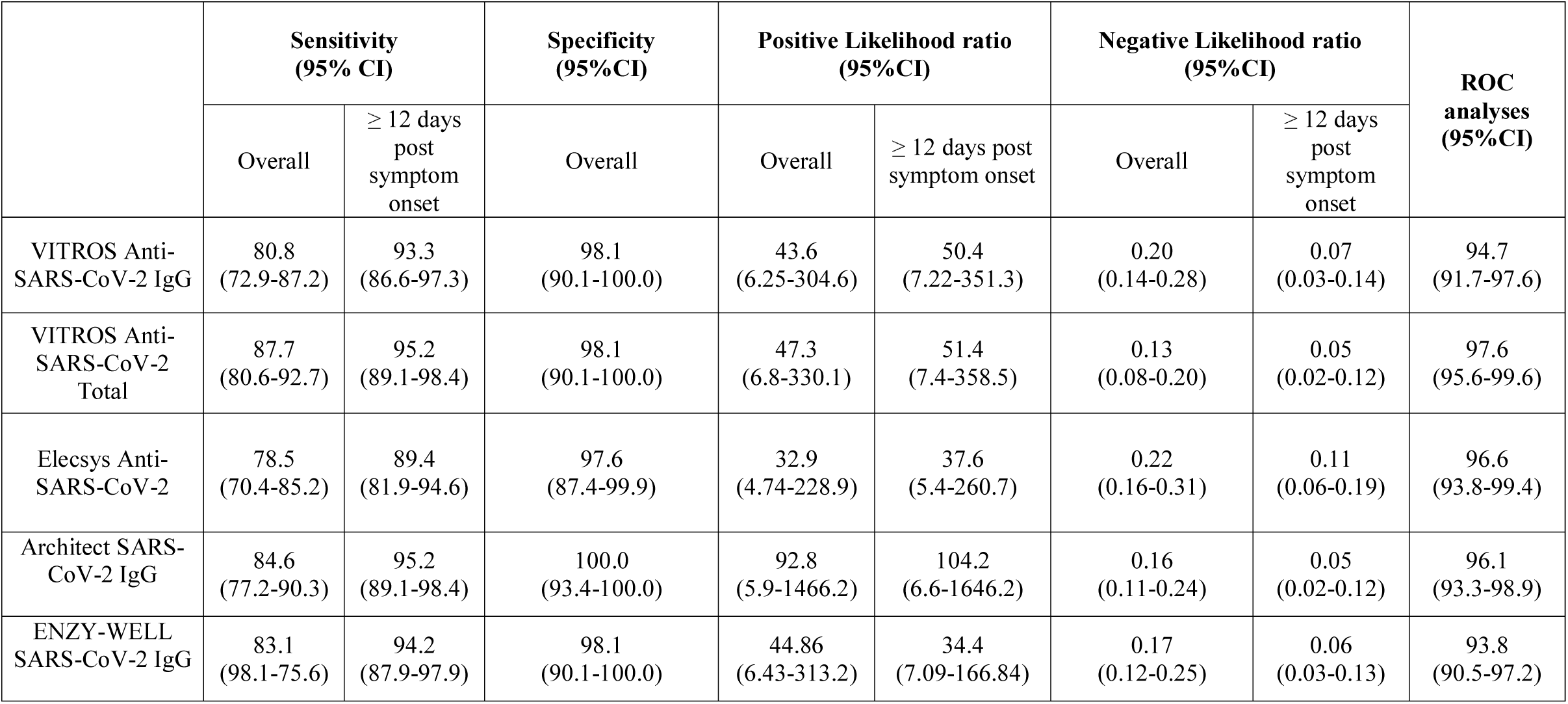
Comparison of the clinical performances of all the studies immunoassays, overall and considering only the period ≥ 12 days post symptoms onset.

### Comparability of immunoassay results

Since the results of serum samples and the corresponding immunoassays’ cut-offs were used to derive either positive or negative test results, pairwise comparisons of all tests by Cohen’s kappa and overall agreements (in percentages) were calculated (Table 5).

**Table 5:** Agreement and Cohen’s kappa of the immunoassays under evaluation.

|  | Immunoassay agreement results |  |  |  |
| --- | --- | --- | --- | --- |
|  | VITROS Anti-SARS-CoV-2 IgG | VITROS Anti-SARS-CoV-2 Total | Elecsys Anti-SARS-CoV-2 | Architect SARS-CoV-2 IgG |
| VITROS Anti-SARS-CoV-2 Total | Agreement = 94.5%<br>Cohen's kappa = 0.886<br>SE = 0.073 | - | - | - |
| Elecsys Anti-SARS-CoV-2 | Agreement = 91.2%<br>Cohen's kappa = 0.817<br>SE = 0.076 | Agreement = 92.4%<br>Cohen's kappa = 0.836<br>SE = 0.075 | - | - |
| Architect SARS-CoV-2 IgG | Agreement = 93.5%<br>Cohen's kappa = 0.865<br>SE = 0.073 | Agreement = 94.5%<br>Cohen's kappa = 0.885<br>SE = 0.074 | Agreement = 92.4%<br>Cohen's kappa = 0.840<br>SE = 0.076 | - |
| ENZY-WELL SARS-CoV-2 IgG | Agreement = 94.0%<br>Cohen's kappa = 0.877<br>SE = 0.073 | Agreement = 97.3%<br>Cohen's kappa = 0.943<br>SE = 0.073 | Agreement = 93.0%<br>Cohen's kappa = 0.852<br>SE = 0.076 | Agreement = 96.2%<br>Cohen's kappa = 0.921<br>SE = 0.073 |

SE = standard error.

### Plaque reduction neutralization test (PRNT)

The signal-to-cut-off (S/CO) ratios of the examined assays, including Liaison SARS-CoV-2 S1/S2 IgG, ENZY-WELL SARS-CoV-2 IgA and IgM and the corresponding PRNT_50_ titers for the 52 tested SARS-CoV-2 serum samples are shown in Figure 2. Overall, positive associations were found between PRNT_50_ titer and S/CO ratios. The highest correlation was obtained with ENZY-WELL SARS-CoV-2 IgM (rho = 0.732, p < 0.001) followed by ENZY-WELL SARS-CoV-2 IgG (rho = 0.541, p < 0.0001), ENZY-WELL SARS-CoV-2 IgA (rho = 0.480, p < 0.001), VITROS Anti-SARS-CoV-2 IgG (rho = 0.368, p < 0.0001), Architect SARS-CoV-2 IgG (rho = 0.347, p = 0.0183), and Liaison SARS-CoV-2 S1/S2 IgG assay (rho = 0.317, p = 0.033). No statistically significant correlations were found for VITROS Anti-SARS-CoV-2 Total (rho = 0.139, p = 0.358) and Elecsys Anti-SARS-CoV-2 (rho = 0.079, p = 0.600). Figure 3 (panel A) shows the distributions of PRNT_50_ titers subdivided by disease severity (Kruskal-Wallis test, chi2 = 9.70, p = 0.0078). Notwithstanding the limited number of samples, asymptomatic/paucisymptomatic cases presented a significantly lower PRNT_50_ titer than moderate/severe cases (Asympt vs. Moderate B-Adj p = 0.019 and Asympt vs. Severe B-Adj p = 0.0065). The relationship between Ab neutralization activity and time post symptom onset (panel B) was found to be statistically significant (rho = −0.345, p = 0.0164). The PRNT_50_ titer did not correlate with age (rho = 0.181, p = 0.192), but was correlated with gender (rho = 0.347, p = 0.011), whilst no significant correlation was found when the analysis was adjusted for disease severity.

**Figure 2:**
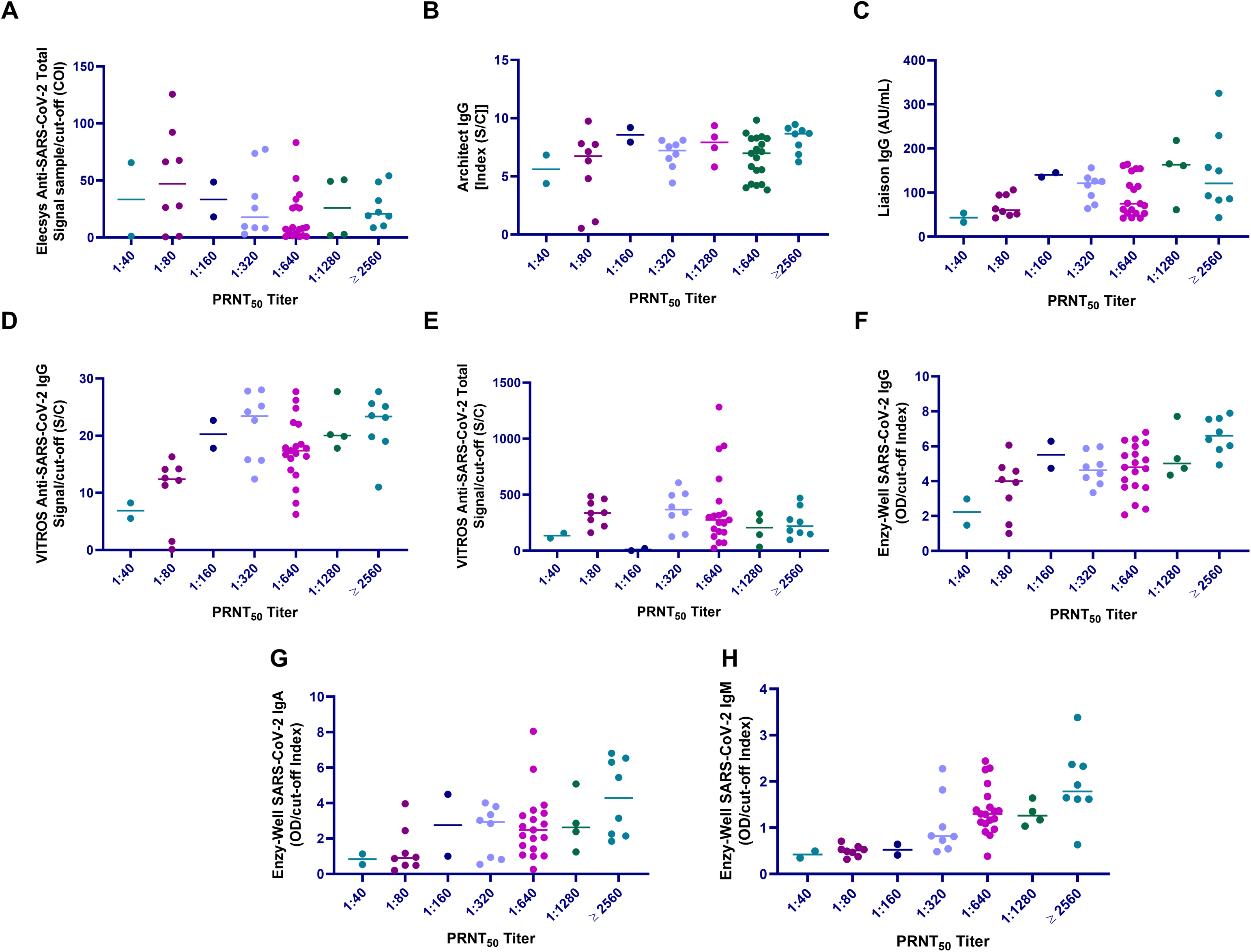
Comparison of plaque reduction neutralization test (PRNT) and immunoassay results. A) Roche Elecsys Anti-SARS-CoV-2 Total against N antigen, B) Abbott Architect Anti-SARS-CoV-2 IgG against N antigen, C) Liaison Diasorin Anti-SARS-CoV-2 IgG Against S1/S2 protein, D) Ortho Clinical Diagnostics VITROS Anti-SARS-CoV-2 IgG against S protein, E) Ortho Clinical Diagnostics VITROS Anti-SARS-CoV-2 Total against S1 protein, F) Enzy-Well, Anti-SARS-CoV-2 IgG against native antigen; G) ENZY-WELL Anti-SARS-CoV-2 IgA against native antigen, H) ENZY-WELL Anti-SARS-CoV-2 IgM against native antigen

**Figure 3:**
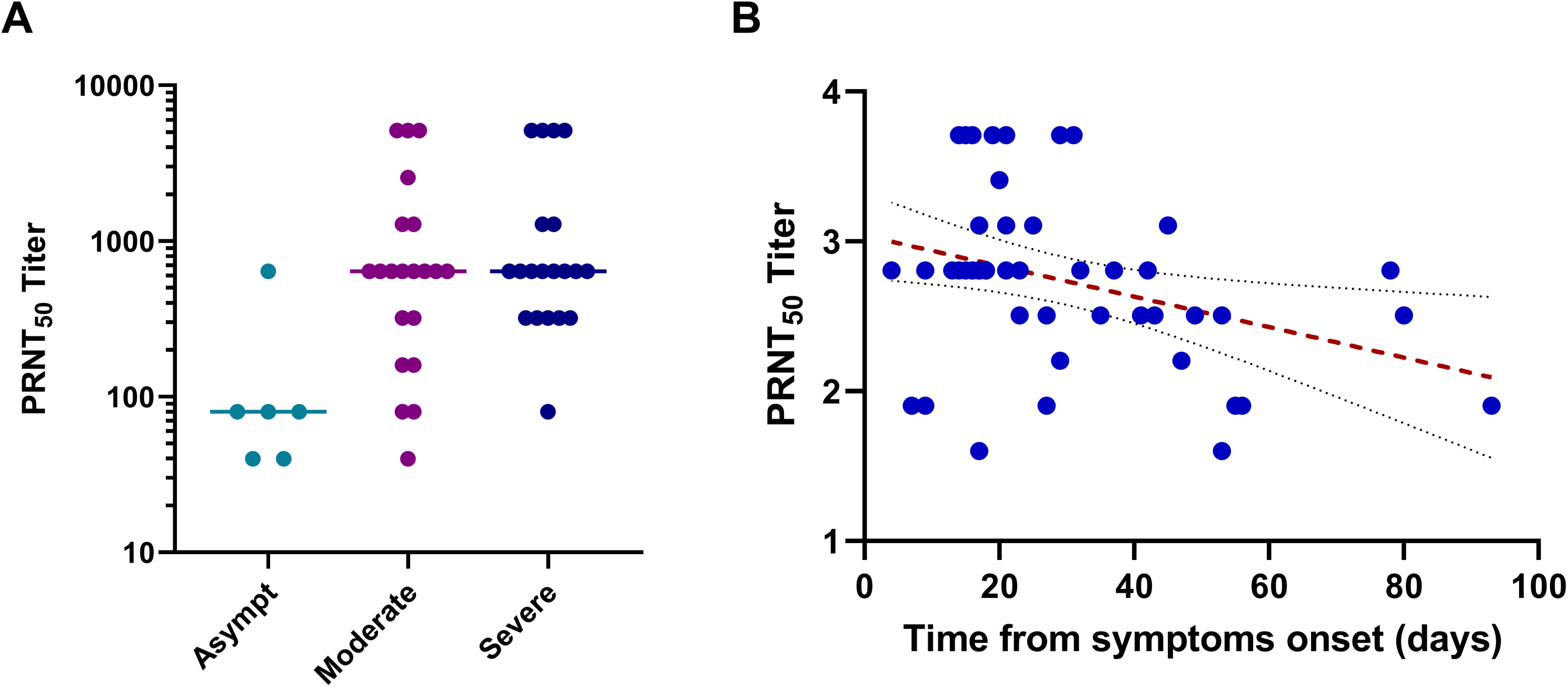
Plaque reduction neutralization test (PRNT) results, disease severity and time from symptoms onset (days). Asympt = asymptomatic/paucisymptomatic SARS-CoV-2 patients; Moderate and Severe = SARS-CoV-2 positive patients without and with air ventilation support, respectively.

## Discussion

In the last few months, numerous SARS-CoV-2 serology assays have been developed. The complexity of COVID-19 has called for careful study design to obtain meaningful information, and some of the assays have not yet been extensively validated by independent laboratories. In a recent meta-analysis, it was pointed out that most studies on SARS-CoV-2 serology have assessed sensitivity without considering time from onset of symptoms and/or including COVID-19-positive cases that are RT-PCR-negative ^13^. Morevoer, researchers are currently facing other knowledge gaps in SARS-CoV-2 serology. For example, the understanding of the neutralization activity of serum antibodies against viral particles is incomplete, calling for the development of strategies to improve our understanding of their relationship with SARS-CoV-2 Ab detected by conventional immunoassays ^14^. Neutralization against SARS-CoV-2 should be evaluated in parallel to serological tests as neutralization ability provides evidence of the mechanism of development of viral immunity and is the gold standard in terms of assay specificity ^15^.

In this retrospective study, the analytical and clinical performances of four commercially available CLIA assays and one ELISA assay (Table 2) have been evaluated and compared with neutralization activity using the plaque reduction neutralization test (PRNT). The neutralization activity was evaluated also with respect Liaison SARS-CoV-2 S1/S2 IgG and ENZY-WELL SARS-CoV-2 IgA and IgM. Before conducting the study, precision at two concentration levels and linearity were assessed for CLIA by using a standardized protocol according to the CLSI EP15-A3 and CLSI EP06-A (Table 3 and Fig. 1) ^9,10^ The results obtained demonstrated that both repeatability and intermediate precisions were comparable with other immunoassays performances for the highest concentration levels ^3,5^, whilst for the lowest levels, less satisfactory results were obtained for VITROS Anti-SARS-CoV-2 Total and Architect SARS-CoV-2 IgG. Linearity was assessed in a range of values covering manufacturers’ cutoffs (Fig. 1). Linear results were obtained for Elecsys Anti-SARS-CoV-2, whereas the performance of other immunoassays was less effective, demonstrating that a double serum antibodies concentration will not correspond to a double in S/CO ratio. To assess the clinical performances of immunoassays, a total of 184 leftover samples obtained from Negative Healthy workers, Autoimmune patients/pregnant women, asymptomatic/paucisymptomatic and Moderate/Severe SARS-CoV-2 patients were evaluated (Table 1). The 21 autoimmune patients and the eight pregnant women, who were SARS-CoV-2 negative, were included in order to evaluate possible analytical interferences. According to the suggestion on study design for SARS-CoV-2 serology, clinical performances were evaluated by considering the total time frame (overall data), and limiting the analyses to sera collected 12 days after the onset of symptoms as this period greatly impacts on immunoassay sensitivity ^5^. Results of ROC analyses showed overlapping performances for all immunoassays (Table 4). VITROS Anti-SARS-CoV-2 Total provided the best results, with an AUC over 97% (lower confidence limit > 95%). Considering overall data, Elecsys Anti-SARS-CoV-2 and Architect SARS-CoV-2 IgG sensitivities obtained in our study are similar those reported by Kohmer et al. and Theel et al. ^16,17^. Sensitivities differed when only samples collected after 12 days post symptom onset were evaluated. This was expected since, after SARS-CoV-2 infection, antibody levels begin to rise as from the second week of onset of symptoms ^18^. Evaluations after 12 days post symptom onset confirmed overall excellent results for all immunoassays, anti-SARS-CoV-2 Total and Architect SARS-CoV-2 IgG performances being the best. However, in contrast with statements in manufacturers’ inserts, our data show that 100% true positive results cannot be obtained, even when the sample collection time is more than 14 days after onset of symptoms. These results are in agreement with findings made by other Authors on comparing immunoassays results ^15,19,20^. Interestingly, asymptomatic/paucisymptomatic SARS-CoV-2 patients were correctly identified as positive by all immunoassays. Differently, 95% CI of specificities reached 100% for all the assays, and these data are in line with manufacturers’ inserts. Although excellent positive- (PLR) and negative- (NLR) likelihood ratios were obtained for all methods, Architect SARS-CoV-2 IgG PLR outperform all other immunoassays. PPV and NPV were further estimated considering two prevalence settings (4% and 10%). The highest achievable PPV and NPV values were 100% and 99.8% for PPV and NPV, respectively, obtained with Architect SARS-CoV-2 IgG and at a prevalence setting of 10%.

To provide insight on neutralization activity compared with immunoassays results, PRNT assay was performed on 52 samples from SARS-CoV-2 positive subjects. With the exception of VITROS Anti-SARS-CoV-2 Total and Elecsys Anti-SARS-CoV-2, other immunoassay results were correlated with PRNT_50_ titer. However, as shown in Figure 3, the dynamic range of measured antibodies, from the lowest to the highest PRNT_50_ titer, is very limited, this being in line with the data reported by Jääskeläinen et al. ^15^. This might be due to several factors, such as: a) antibodies measurements being specifically developed for detecting positive/negative subjects, improving RT-PCR diagnosis of COVID-19 or b) there being a non-linear response between the real antibody concentration and instrumental signals. Notably, results of Liaison SARS-CoV-2 S1/S2 IgG assay were similar to those obtained with other immunoassays. At correlation analysis, no differences could be detected on comparing immunoassays developed against Spike or Nucleocapsid proteins. Differently, the strongest correlation with PRNT_50_ titer was found for ENZY-WELL ELISA results, developed with a native antigen of SARS-CoV-2 (rho = 0.541, p < 0.001). Recently, Perera et al. found a slightly stronger correlation (rho = 0.67) between plaque reduction neutralization results and an in-house developed IgG ELISA with recombinant RBD of the spike protein as coated antigen ^21^. In addition, we found a highly significant correlation between PRNT and IgM Elisa results, thus confirming the data reported by Perera et al. ^21^. Another report, which compare IgG or total antibodies measurement of three ELISA, two CLIA and two lateral flow tests, in a total of 100 SARS-CoV-2 convalescent plasma donors, found a good correlation (rho > 0.700) between ELISA (Euroimmun IgG and Wantai Total antibodies) and neutralization titer ^22^.

In asymptomatic/paucisymptomatics, PRNT_50_ titer values were lower than in moderate/severe SARS-CoV-2 patients. Furthermore, a significant negative correlation was found between the PRNT_50_ titer and the time interval from symptom onset (Figure 3).

The present paper has limitations: first, neutralizing antibodies were tested in a limited number of samples the procedure being very complex; second, COVID-19 positive patients were selected retrospectively on the basis of available leftover samples; therefore NPV and PPV could be overestimated. Finally, the relationship between IgM antibodies and neutralizing activity should be studied further in a larger series of patients.

In conclusion, although the performances of SARS-CoV-2 antibody immunoassays are of analytical and clinical value, they could be enhanced by considering the test purposes, emphasizing sensitivity in the screening and specificity in the second-line testing. In addition, a further search should be made for a better dynamic range and a stronger correlation with respect to antibody neutralization activity, in order to, above all, obtain information needed for effective passive antibody therapy and vaccine development against SARS-CoV-2 virus.

## Data Availability

All data have been collected and available in saved computer facilities and eventually some raw data should be requested, if needed

## Acknowledgments

We thank Daniela Rinaldi (medical laboratory scientists) for their valuable technical support. We acknowledge Abbott Laboratories, Diesse Diagnostica Senese, Diasorin, Ortho Clinical Diagnostics, Roche Diagnostic for kindly supplying reagents without any influence in study design and data analysis.

## Author contributions

Study design: MP; AP.

Sample collection and experimental set-up: DN; SZ; CC; FB; MPag; AB.

Data Collection and statistical analyses: DN; DB; AP.

Writing of the manuscript: AP; LS; MP.

Review of the manuscript: FB; AP; LS; MP.

